# The psychological distress and coping styles in the early stages of the 2019 coronavirus disease (COVID-19) epidemic in the general mainland Chinese population: a web-based survey

**DOI:** 10.1101/2020.03.27.20045807

**Authors:** Hui-yao Wang, Qian Xia, Zhen-zhen Xiong, Zhi-xiong Li, Wei-yi Xiang, Yi-wen Yuan, Ya-ya Liu, Zhe Li

## Abstract

**Background:** As the epidemic outbreak of 2019 coronavirus disease (COVID-19), general population may experience psychological distress. Evidence has suggested that negative coping styles may be related to subsequent mental illness. Therefore, we investigate the general population’s psychological distress and coping styles in the early stages of the COVID-19 outbreak.

**Methods:** A cross-sectional battery of surveys was conducted from February 1-4, 2020. The Kessler 6 psychological distress scale, the simplified coping style questionnaire and a general information questionnaire were administered on-line to a convenience sample of 1599 in China. Spearman’s correlation was used to measure the correlations among category variables.

**Results:** General population’s psychological distress were significant differences based on age, marriage, epidemic contact characteristics, concern with media reports, and perceived impacts of the epidemic outbreak (all *p* <0.001) except gender (*p*=0.316). Those with a history of visiting Wuhan and a history of epidemics occurring in the community, more concern with media reports, perceived more severe impacts and negative coping style had a higher level of psychological distress, which was significantly positively correlated with a history of visiting Wuhan (r=0.548, *p*<0.001), a history of epidemics occurring in the community (r=0.219, *p*<0.001), and concern with media reports (r=0.192, *p*<0.001). Coping styles were significantly different across all category variables (all p <0.001), and negatively correlated with other category variables (all *p*<0.01) except age and marriage. Psychological distress was significantly negatively correlated with the coping style (r=-0.573, *p*<0.01).

**Conclusions:** In the early stages of COVID-19, general population with epidemic contact characteristics, excessive concern with media reports, and perceived more severe impacts have higher levels of psychological distress. Psychological distress was significantly negatively correlated with the coping style. Interventions should be implemented early, especially for those population with a high level of psychological distress and/or with a negative coping style.

## Introduction

The epidemic of the 2019 coronavirus disease (COVID-19) has aroused widespread concern throughout society in China. Because the virus can be transmitted through droplets, contact, etc. [1], cities in many regions of China have closed non-essential public places, restricted mass gathering activities, and enacted other control measures to effectively control the spread of the virus [2]. The epidemic has had a strong impact on general population’s daily life. At the same time, as the epidemic continues, general population gradually experience different levels of psychological distress, such as nervousness, fear of infection, anxiety, depression, sleep problems, and inattention [3,4]. Previous studies have reported that some psychological problems often occur during similar epidemic [5,6] or other traumatic stress events, such as natural disasters [7,8], disease [9], or long-term employment in high stress occupations [10-12], and may last for a long time [13,14].

When faced with stress or traumatic experiences, general population often responds differently, with some responding positively and others responding negatively. Evidence has suggested that coping styles in the face of stress have an impact on the quality of general population’s life [15,16], and negative coping styles may be related to psychological distress or mental illness such as post-traumatic stress disorder (PTSD), anxiety, and depression [7,8,12]. For this reason, we conducted this study in the early stages of this epidemic to investigate the general population’s psychological distress and coping style related to the epidemic of COVID-19 so that those who have high levels of psychological distress and/or respond negatively can be detected early and undergo timely intervention.

## Methods

This study was conducted through an online survey, starting at 16:00 on February 1, 2020 and ending at 24:00 on February 4, and the survey was approved by the ethical review board of the West China Hospital of Sichuan University. The snowball sampling method was used to invite subjects. All invitees completed the questionnaire online via Questionnaire Star (https://www.wjx.cn). An initial set of invitees (10 participants) was chosen to ensure broad representation of age, gender, occupation, education level, and city. This set of invitees then forwarded the questionnaire to 10 companions whom they considered suitable for the survey, and this second set forwarded the questionnaire in the same way. The study included a general population aged 18 years or older who volunteered to participate in the study, and respondents were excluded if they reported a history of mental illness and/or could not complete the online survey independently.

### Data collection

A self-made questionnaire was used to collect demographic and epidemiological information of participants, including gender, age, marriage, epidemic contact and concern characteristics, and perceived epidemic impacts of the epidemic of COVID-19.

### Psychological distress assessment

The Kessler 6 psychological distress Scale (K6) was used to assess the psychological distress of participants; this scale has been proven to have cross-cultural reliability and validity [17]. It contains six questions that ask participants to rate how often they have felt ‘nervous’, ‘hopeless’, ‘restless or fidgety’, ‘so depressed that nothing could cheer you up’, ‘that everything was an effort’, and ‘worthless’ during the past 30 days.

### Coping style assessment

The Simplified Coping Style Questionnaire (SCSQ) was used to assess the participants’ coping styles during the COVID-19 epidemic; this questionnaire has been proven to have good reliability and validity in Chinese [18]. The SCSQ contains twenty items, with each item using a four-point score (0□=□never, 1□=□seldom, 2□=□often, 3□=□always), and two subscales: positive coping (12 items) and negative coping (8 items). According to the average and standard deviation of the positive coping style and the negative coping style scores, a Z conversion is used to calculate their respective standard scores, and then, the negative coping standard scores are subtracted from the positive coping style standard scores to calculate the tendency value of coping style. A result greater than 0 was defined as a participant adopting a positive coping style when faced with stress, and a result less than 0 was defined as a participant adopting a negative coping style [19].

### Statistical analysis

Differences in psychological distress (K6 score) among categorical variables were tested by t-tests or one-way analysis of variance, and differences in coping styles were tested by chi-squared tests. Correlations among all categorical variables were measured by Spearman’s correlation. All statistical analyses were conducted in SPSS version 22.0 (IBM, Chicago, IL, USA), and p <0.05 was considered to be statistically significant.

### Quality control

The same IP address could be used only once to complete the questionnaire, which did not collect any personal information such as names, thereby ensuring anonymity and honest responses. The time spent on each questionnaire was monitored automatically, and the whole questionnaires completed in fewer than 120 seconds were rejected as invalid.

## Results

### Sample characteristics

There were 1607 individuals from 26 regions of China who completed this survey, and 1599 (99.5%) were included in the analysis participants. Among all participants, 1068 were female (66.8%); ages ranged from 18 to 84 years old (mean 33.9±12.3 years); 914 were married (57.2%); 326 had a history of visiting Wuhan (20.4%); and 333 had a history of epidemics occurring in their community (20.8%) (Table 1).

**Table 1.** Sample description

| Variables | n (%) |
| --- | --- |
| **Total** | 1599(100.0) |
| **Demographic characteristics** |  |
| Gender |  |
| female | 1068(66.8) |
| male | 531(33.2) |
| Age (mean $33.9 \pm 12.3$, years) | |
| 18-30 | 722(45.2) |
| 31-40 | 471(29.5) |
| 41-50 | 254(15.9) |
| >50 | 152(9.5) |
| Marriage |  |
| single | 624(39.0) |
| married | 914(57.2) |
| divorced | 61(3.8) |
| **Epidemic contact characteristics** |  |
| History of visiting Wuhan |  |
| no | 1273(79.6) |
| yes | 326(20.4) |
| History of epidemics occurring in the community |  |
| no | 1266(79.2) |
| yes | 333(20.8) |
| **Concern with media reports related to the epidemic** |  |
| less concerned | 16(1.0) |
| concerned | 141(8.8) |
| more concerned | 428(26.8) |
| extremely concerned | 1014(63.4) |
| **Perceived impacts of the epidemic** |  |
| Changes over living situations |  |
| feel relax | 209(13.1) |
| no change | 479(30.0) |
| feel nervous | 911(56.9) |
| Emotional control |  |
| no difficult | 832(52.0) |
| less difficult | 304(19.0) |
| difficult | 70(4.4) |
| more difficult | 70(4.4) |
| extremely difficult | 323(20.2) |
| Epidemic-related dreams |  |
| no | 987(61.7) |
| less | 151(9.4) |
| general | 115(7.2) |
| more | 33(2.1) |
| extremely large | 313(19.6) |

### Comparison of the psychological distress

The results revealed significant differences in the participants’ psychological distress based on age, marriage, epidemic contact characteristics, concern with media reports related to the epidemic, and perceived impacts of the epidemic (all *p* <0.001); there were no significant differences based on gender (*p* =0.316). As age increases and marital status changes, K6 scores have a downward trend. Those with a history of visiting Wuhan and a history of epidemics occurring in the community have a higher level of psychological distress than those without such experiences. The psychological distress tended to increase with concern with media reports related to the epidemic and perceived impacts of the epidemic. At the same time, the results also show that those with a negative coping style have a higher level of psychological distress than those with a positive coping style (Table 2).

**Table 2.**
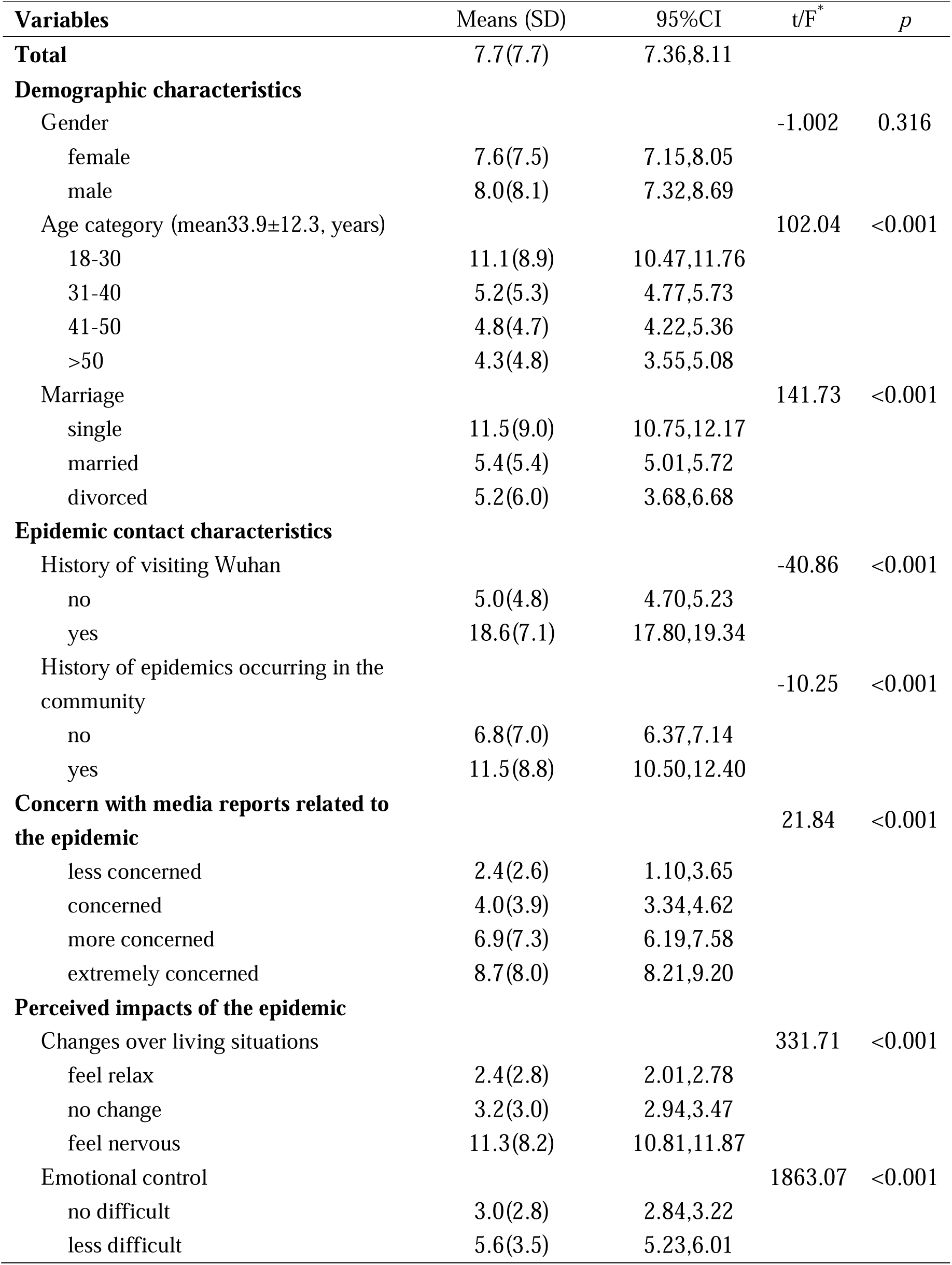
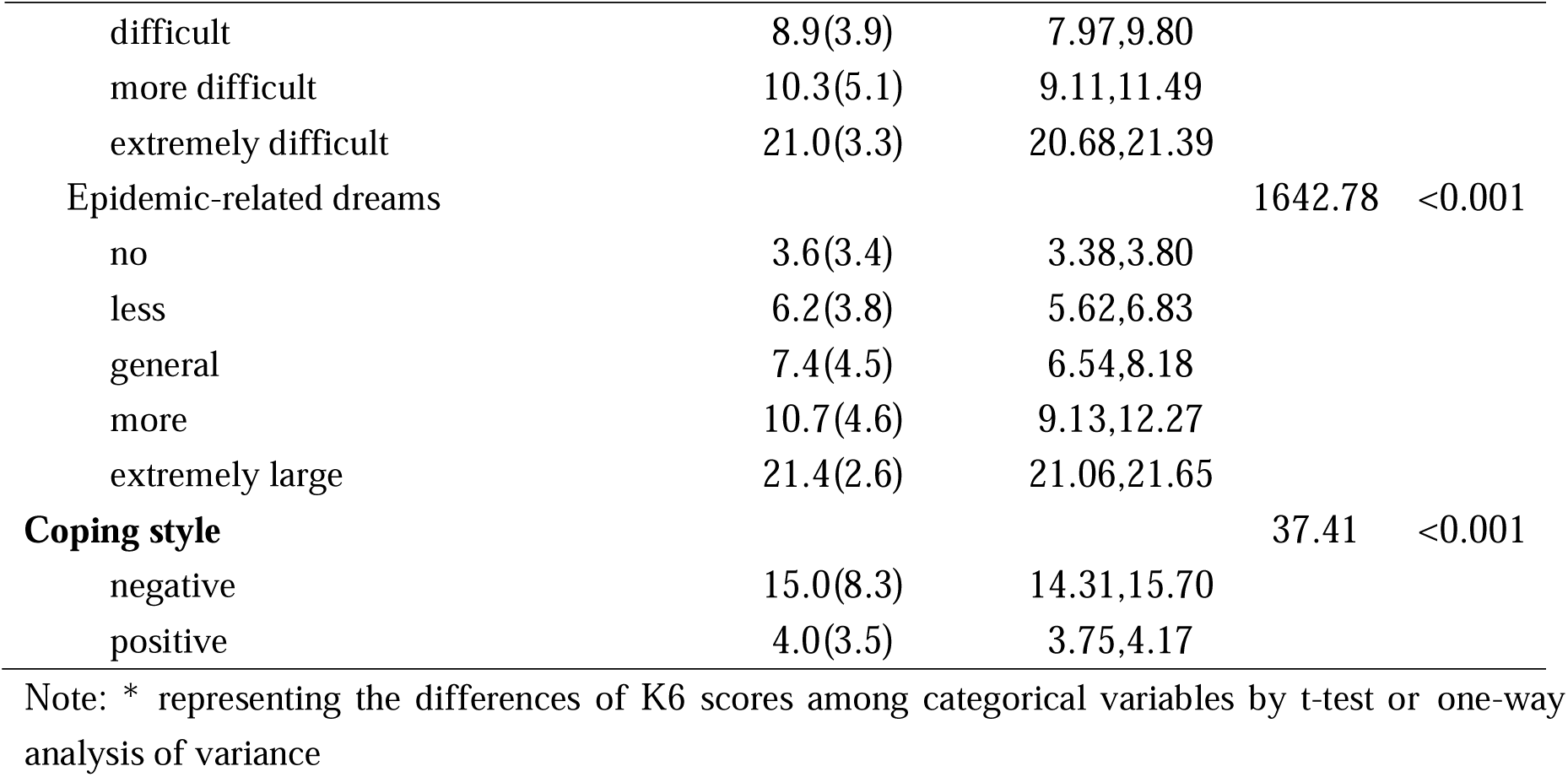
Psychological distress (K6 scores) of participants (n=1599).

| Variables | Means (SD) | 95%CI | t/F* | p |
| --- | --- | --- | --- | --- |
| **Total** | 7.7(7.7) | 7.36,8.11 |  |  |
| **Demographic characteristics** |  |  |  |  |
| Gender |  |  | -1.002 | 0.316 |
| female | 7.6(7.5) | 7.15,8.05 |  |  |
| male | 8.0(8.1) | 7.32,8.69 |  |  |
| Age category (mean33.9±12.3, years) |  |  | 102.04 | <0.001 |
| 18-30 | 11.1(8.9) | 10.47,11.76 |  |  |
| 31-40 | 5.2(5.3) | 4.77,5.73 |  |  |
| 41-50 | 4.8(4.7) | 4.22,5.36 |  |  |
| >50 | 4.3(4.8) | 3.55,5.08 |  |  |
| Marriage |  |  | 141.73 | <0.001 |
| single | 11.5(9.0) | 10.75,12.17 |  |  |
| married | 5.4(5.4) | 5.01,5.72 |  |  |
| divorced | 5.2(6.0) | 3.68,6.68 |  |  |
| **Epidemic contact characteristics** |  |  |  |  |
| History of visiting Wuhan |  |  | -40.86 | <0.001 |
| no | 5.0(4.8) | 4.70,5.23 |  |  |
| yes | 18.6(7.1) | 17.80,19.34 |  |  |
| History of epidemics occurring in the community |  |  | -10.25 | <0.001 |
| no | 6.8(7.0) | 6.37,7.14 |  |  |
| yes | 11.5(8.8) | 10.50,12.40 |  |  |
| **Concern with media reports related to the epidemic** |  |  | 21.84 | <0.001 |
| less concerned | 2.4(2.6) | 1.10,3.65 |  |  |
| concerned | 4.0(3.9) | 3.34,4.62 |  |  |
| more concerned | 6.9(7.3) | 6.19,7.58 |  |  |
| extremely concerned | 8.7(8.0) | 8.21,9.20 |  |  |
| **Perceived impacts of the epidemic** |  |  |  |  |
| Changes over living situations |  |  | 331.71 | <0.001 |
| feel relax | 2.4(2.8) | 2.01,2.78 |  |  |
| no change | 3.2(3.0) | 2.94,3.47 |  |  |
| feel nervous | 11.3(8.2) | 10.81,11.87 |  |  |
| Emotional control |  |  | 1863.07 | <0.001 |
| no difficult | 3.0(2.8) | 2.84,3.22 |  |  |
| less difficult | 5.6(3.5) | 5.23,6.01 |  |  |
| difficult | 8.9(3.9) | 7.97,9.80 |  |  |
| more difficult | 10.3(5.1) | 9.11,11.49 |  |  |
| extremely difficult | 21.0(3.3) | 20.68,21.39 |  |  |
| Epidemic-related dreams |  |  | 1642.78 | <0.001 |
| no | 3.6(3.4) | 3.38,3.80 |  |  |
| less | 6.2(3.8) | 5.62,6.83 |  |  |
| general | 7.4(4.5) | 6.54,8.18 |  |  |
| more | 10.7(4.6) | 9.13,12.27 |  |  |
| extremely large | 21.4(2.6) | 21.06,21.65 |  |  |
| **Coping style** |  |  | 37.41 | <0.001 |
| negative | 15.0(8.3) | 14.31,15.70 |  |  |
| positive | 4.0(3.5) | 3.75,4.17 |  |  |

Note: \* representing the differences of K6 scores among categorical variables by t-test or one-way analysis of variance

### Comparison of the coping style

The ratios of the coping styles are 34.2% negative and 65.8% positive. The comparison found that the coping style was significantly different across all category groups (all *p* <0.001) (Table 3). Compared with the positive coping style, the negative coping style occurred more in those between 18 and 30 years old, single people, those with a history of visiting Wuhan, those with a history of epidemics occurring in the community, those who reported that perceived emotional control was extremely difficult and those who reported having many epidemic-related dreams (Table 3).

**Table 3.**
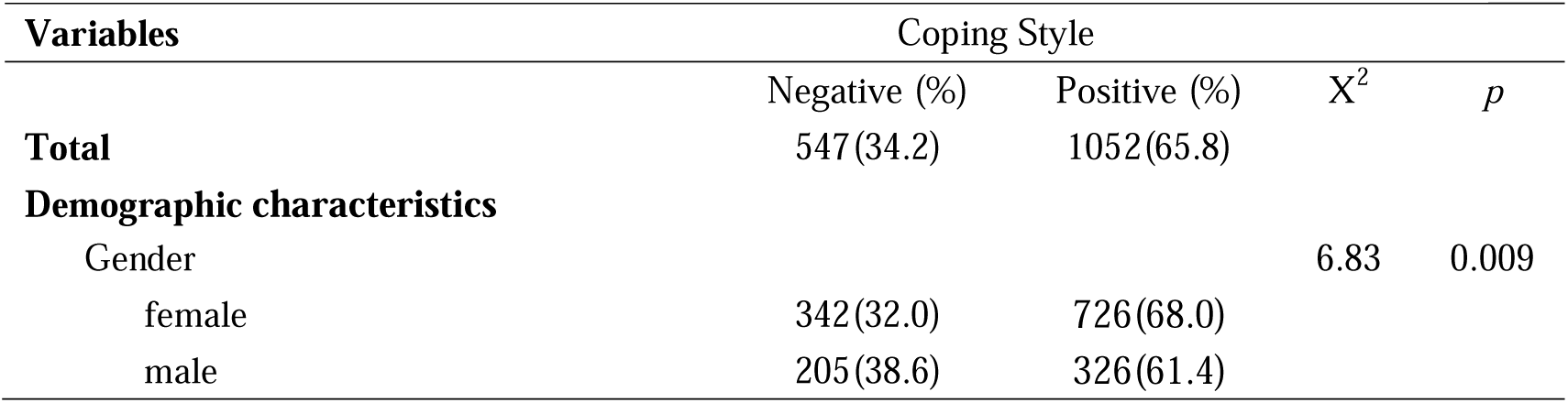
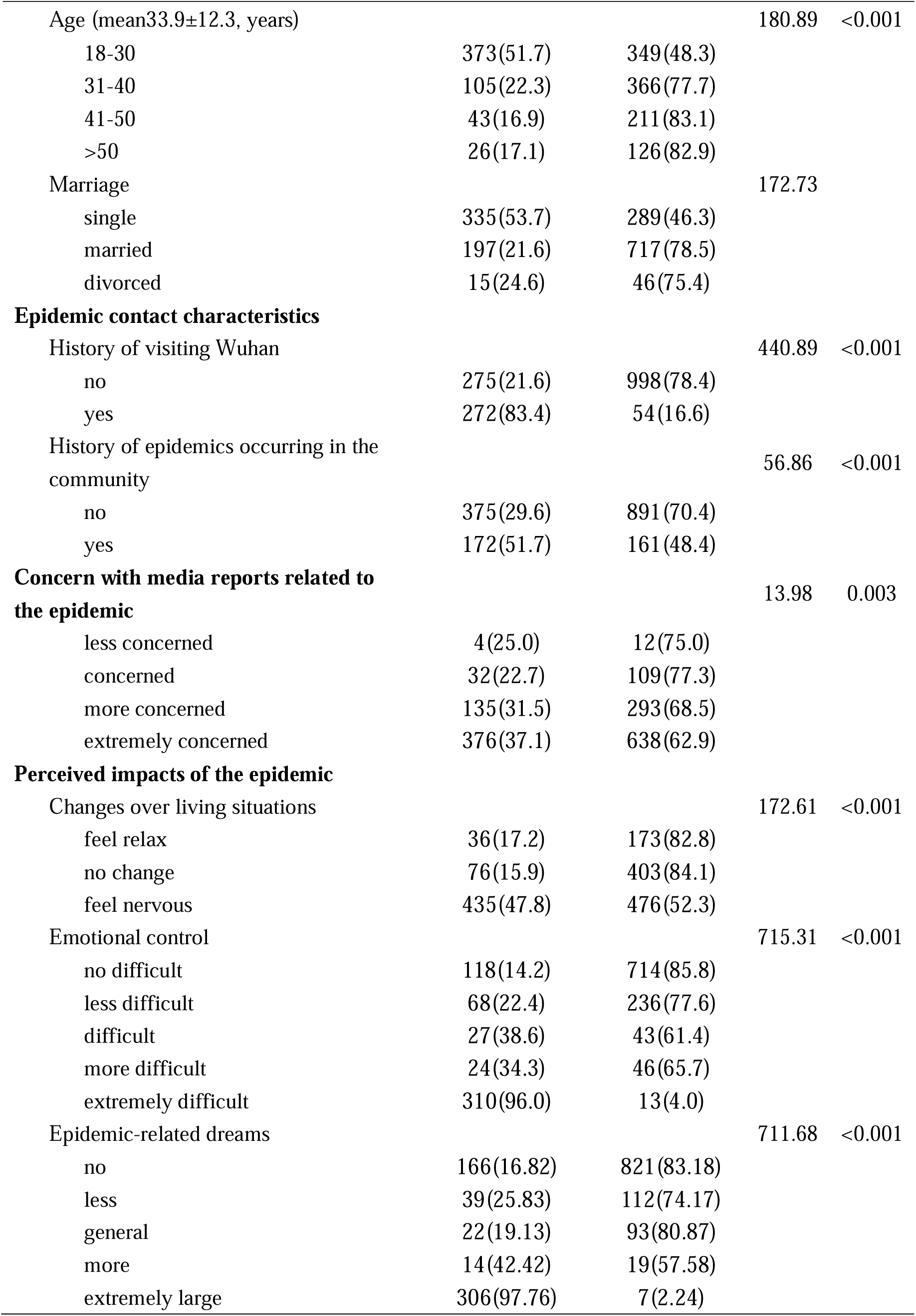
Coping style of participants (n=1599).

### Correlation analysis

The correlation analysis found that psychological distress was significantly positively correlated with a history of visiting Wuhan (r=0.548, *p*<0.001), a history of epidemics occurring in the community (r=0.219, *p*<0.001), concern with media reports related to the epidemic (r=0.192, *p*<0.001), and perceived changes over living situations (r=0.571, *p*<0.001), emotional control (r=0.752, *p*<0.001) and epidemic-related dreams (r=0.708, *p*<0.001). Psychological distress was significantly negatively correlated with age (r=-0.371, *p*<0.001), marriage (r=-301, *p*<0.001) and coping styles (r=-0.573, *p*<0.001); there was no significant correlation with gender. The coping style was significantly positively correlated with age and marriage, and significantly negatively correlated with other category variables. All epidemic contact characteristics and concern with media reports related to the epidemic were significantly positively correlated with perceived impacts of the epidemic (Table 4).

**Table 4.**
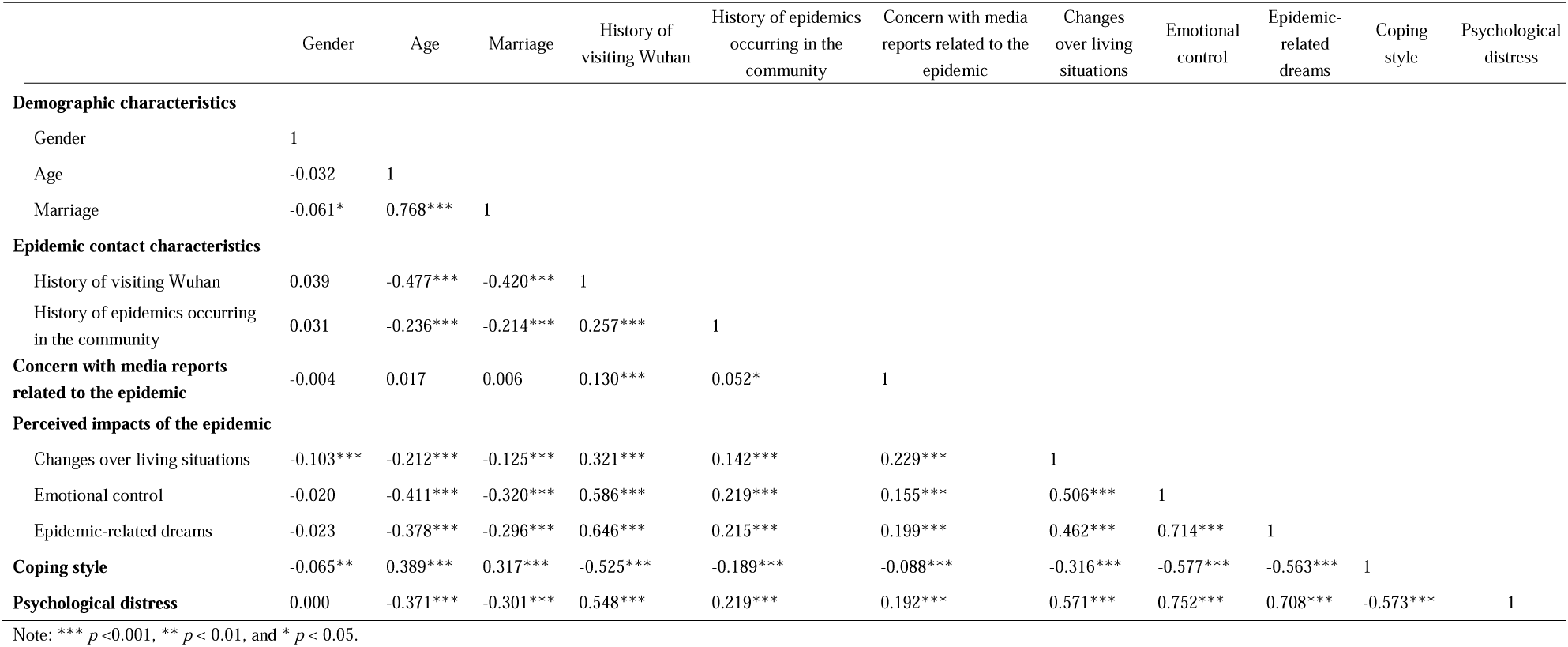
Correlations of demographic, epidemic contact and concern characteristics, perceived epidemic impacts, coping style and psychological distress in all participants(n=1599) (Spearman’s rank correlation)

|  |  |  |  |  |  |  |  |  |  |  |  |
| --- | --- | --- | --- | --- | --- | --- | --- | --- | --- | --- | --- |
|  | Gender | Age | Marriage | History of visiting Wuhan | History of epidemics occurring in the community | Concern with media reports related to the epidemic | Changes over living situations | Emotional control | Epidemic-related dreams | Coping style | Psychological distress |
| **Demographic characteristics** |  |  |  |  |  |  |  |  |  |  |  |
| Gender | 1 |  |  |  |  |  |  |  |  |  |  |
| Age | -0.032 | 1 |  |  |  |  |  |  |  |  |  |
| Marriage | -0.061* | 0.768*** | 1 |  |  |  |  |  |  |  |  |
| **Epidemic contact characteristics** |  |  |  |  |  |  |  |  |  |  |  |
| History of visiting Wuhan | 0.039 | -0.477*** | -0.420*** | 1 |  |  |  |  |  |  |  |
| History of epidemics occurring in the community | 0.031 | -0.236*** | -0.214*** | 0.257*** | 1 |  |  |  |  |  |  |
| **Concern with media reports related to the epidemic** | -0.004 | 0.017 | 0.006 | 0.130*** | 0.052* | 1 |  |  |  |  |  |
| **Perceived impacts of the epidemic** |  |  |  |  |  |  |  |  |  |  |  |
| Changes over living situations | -0.103*** | -0.212*** | -0.125*** | 0.321*** | 0.142*** | 0.229*** | 1 |  |  |  |  |
| Emotional control | -0.020 | -0.411*** | -0.320*** | 0.586*** | 0.219*** | 0.155*** | 0.506*** | 1 |  |  |  |
| Epidemic-related dreams | -0.023 | -0.378*** | -0.296*** | 0.646*** | 0.215*** | 0.199*** | 0.462*** | 0.714*** | 1 |  |  |
| **Coping style** | -0.065** | 0.389*** | 0.317*** | -0.525*** | -0.189*** | -0.088*** | -0.316*** | -0.577*** | -0.563*** | 1 |  |
| **Psychological distress** | 0.000 | -0.371*** | -0.301*** | 0.548*** | 0.219*** | 0.192*** | 0.571*** | 0.752*** | 0.708*** | -0.573*** | 1 |

Note: \*\*\* $p < 0.001$ , \*\* $p < 0.01$ , and \* $p < 0.05$ .

## Discussion

The results of the study suggest that the general population with a history of visiting Wuhan, those with a history of epidemics occurring in their community, and those who perceived more severe impacts of the epidemic of COVID-19 on their living situations, emotional control, and epidemic-related dreams have a higher level of psychological distress than those with none or little of these experiences. These findings are consistent with the findings of previous studies. The traumatic stress experience during the occurrence of emergency events, such as major public events or natural disasters, is often related to the general population’s psychological distress and subsequent mental illness [5,6,8].

Furthermore, the study found that there were more participants who exhibited a negative coping style (34.2%), especially among younger, single, those with a history of epidemic contact, and those who perceived severe impacts of the epidemic of COVID-19.

Other studies related to traumatic stress events have reported that those in the general population with traumatic stress experiences were more likely to adopt a negative coping style [9]. Some studies have shown that this phenomenon is related to the general population’s brain functional connectivity [15], while others have shown that it is related to social support [20,21]. The underlying mechanism of this association has yet to be elucidated, which may be the result of the combined effect of the two mechanisms. Further study is needed to confirm this hypothesis. More importantly, the study also found that the general population with negative coping styles has higher psychological distress than those with positive coping styles. Many previous studies have shown that different coping styles, especially negative coping styles, for trauma stress events are usually related to subsequent mental illness [7,10,22]. Therefore, this part of the general population with a negative coping style should be given attention and follow-up, and appropriate interventions should be given if necessary.

Finally, this study included concern with media reports related to the epidemic of COVID-19 in the analysis. Previous studies have reported that post-disaster mental health problems, such as post-traumatic stress disorder (PTSD), may be related to media reports in addition to direct exposure to the disaster environment [23,24]. The study found that with increasing concern with media reports related to epidemic, the general population’s psychological distress level has also increased accordingly; this study also found that the general population’s coping style was negatively related to concern with media reports. This result suggested that during the epidemic of COVID-19, the degree of concern with media reports affected the general population’s psychological distress level and coping style, which might be related to that media reports could affect the general population’s perceptions of the disease and what preventive measures they would take [25,26]; further research is needed to confirm this relation.

In conclusion, this study found that the epidemic of COVID-19 caused different levels of psychological distress. Those with epidemic contact characteristics, excessive concern with media reports, and those who perceived more impacts of the epidemic reported higher levels of psychological distress. The psychological distress was significantly different in general population and was significantly negatively correlated with coping style. Therefore, appropriate interventions should be implemented early to address the impacts of such epidemics, especially for those in the general population with a high level of psychological distress and/or with a negative coping style.

## Limitations

There are several limitations in our study. First, the survey method is based on network invitation rather than face-to-face random sampling, and participants need to be able to use network tools. As a result, the status of the general population who cannot use network tools is unclear. Second, we did not assess whether and how respondents were engaging in prevention. Finally, our study design is cross-sectional and thus cannot capture changes in psychological distress and its predictors over the course of the COVID-19.

## Data Availability

Data cannot be shared publicly because of ethical restrictions.

## Acknowledgments

We thank the general population who participated in this survey and bravely resisted the COVID-19 epidemic, and thank the Questionnaire Star (https://www.wjx.cn) for providing us with a data survey platform.

## Author Contributions

**Data curation**: Zhen-zhen Xiong, Wei-yi Xiang, Zhe Li.

**Formal analysis**: Hui-yao Wang, Qian Xia.

**Investigation**: Zhen-zhen Xiong, Zhi-xiong Li, Wei-yi Xiang, Yi-wen Yuan.

**Validation**: Zhi-xiong Li, Ya-ya Liu, Yi-wen Yuan.

**Writing – original draft**: Hui-yao Wang, Qian Xia.

**Writing – review & editing**: Zhe Li.

